# Risk-targeted interventions for measles control-Lessons from an emergency response project in the Katanga Region, Democratic Republic of the Congo (2021-2023)

**DOI:** 10.64898/2026.07.23.26358765

**Authors:** Birgit Nikolay, Halidou Salou, Charles Matungulu, Gisèle Mbuyi, Noella Mosala, Dominique Muteba, Catherine Eisenhauer, Jean Marie Kafwembe, Patrick Banza Mpiong, Francis Kambol, Wilma Lwabola Numbi, Jacques Muzinga, Steve Ahuka, Elisabeth Pukuta Simbu, Virginie Napolitano, Victor Dara, Matthew J Ferrari, Corinne Danet, Leon Salumu, Etienne Gignoux, Christopher Mambula, Klaudia Porten, Raymond Adeango Adhaku, Aimé Cikomola, Nicole Lubanda, Isabel Amoros Quiles

## Abstract

The Urgepi project is a measles emergency initiative implemented by Médecins Sans Frontières Operational Centre Paris (MSF-OCP) in the Katanga region of the Democratic Republic of Congo, a setting characterized by recurrent measles epidemics with substantial public health impact. To maximize impact, the project employs a risk-targeted approach directing resources to selected health zones to strengthen surveillance, implement preventive vaccination, and ensure timely epidemic response.

This study presents the Urgepi approach, and, based on surveillance data from 2021-2023, evaluates the effectiveness of targeted preventive activities, explores strategies for prioritizing interventions amid multiple alerts, and assesses the overall impact of interventions on measles epidemics.

Preventive vaccination successfully averted epidemics in six of the nine vaccinated high-risk health zones, while nine of ten non-vaccinated priority health zones experienced large epidemics. In the three vaccinated health zones that experienced epidemics, more locally adapted vaccination strategies would have been required to achieve higher coverage. Regarding the project’s approach to prioritize alerts for investigations and interventions, we found that the Urgepi alert prioritization score was positively associated with subsequent epidemic size, suggesting that this approach allowed effective and timely targeting of larger local measles epidemics. Although no clear impact of epidemic response vaccination on case numbers was observed, likely due to biases introduced in case reporting through provision of free case management, a 50% reduction in the median epidemic duration was recorded.

In conclusion, the findings demonstrate the effectiveness of the Urgepi project’s risk-targeted approach for measles prevention and control and highlight the importance of timely and context-specific interventions.

**Key questions:**

- **What is already known on this topic** – Measles can, in theory, be effectively controlled through preventive vaccination or rapid immunization responses during outbreaks. However, many countries continue to face significant operational challenges.
- **What this study adds** – We demonstrate how a risk-targeted approach, with prioritization of both preventive and response actions, can optimize resources in the fight against measles.
- **How this study might affect research, practice or policy** – Similar risk-targeted approaches can help national programs prioritize geographic areas for measles prevention and outbreak response when it is not feasible to cover all regions due to operational constraints.

## Introduction

Measles is a highly contagious viral disease. It remains an important cause of death among young children globally, despite the availability of a safe and effective vaccine. While vaccination has drastically reduced global measles deaths, with an estimated 60 million deaths averted between 2000 and 2023, measles is still common in many developing countries, particularly in parts of Africa and Asia [1].

One of these countries is the Democratic Republic of the Congo (DRC), where a resurgence of measles cases has been observed since 2010 with measles epidemics occurring periodically every 2-3 years despite regular nationwide supplementary vaccination activities. In 2018-2020 the DRC faced an unprecedented nationwide measles epidemic, affecting all 26 provinces with more than 457 000 reported cases and over 8100 deaths and was followed by another nationwide epidemic of similar scale during 2021-2023 with more than 512 000 cases and 8400 deaths [2,3]. These numbers likely represent only a fraction of the actual cases and deaths that occurred in the community [4–6].

Médecins Sans Frontières Operational Centre Paris (MSF-OCP) has a long-standing history of supporting measles epidemic responses in DRC [4,7–10]. Despite this extensive experience, delivering timely and effective responses to measles outbreaks has remained a persistent operational challenge. To strengthen strategies for both the prevention and control of measles epidemics in the DRC and beyond, MSF-OCP, in collaboration with the DRC Ministry of Health (MoH), launched a measles emergency response project in 2018.

The “Urgepi” project is based in the Katanga region, covering the provinces of Haut Katanga, Haut Lomami, Lualaba, and Tanganyika, one of the areas most heavily affected by measles in the country. The project encompasses a comprehensive set of activities aimed at improving measles prevention and outbreak response through four key components: (i) Preventive vaccination in high-risk health zones during non-epidemic periods; (ii) Strengthened surveillance and laboratory confirmation of suspected cases to enable earlier detection and faster confirmation of outbreaks; (iii) Epidemic response interventions, including case management and reactive vaccination campaigns to reduce morbidity and mortality [11]; and (iv) Operational research to develop, adapt, and evaluate tools and strategies for measles control.

To maximize the impact of MSF-OCP’s operational capacity against measles in the Katanga region, an area encompassing 68 health zones and a population of over 15 million people [12], the Urgepi project employs a risk-targeted approach. Guided by epidemiological evidence, this strategy prioritizes health zones for surveillance, investigation, and intervention, allowing for more efficient allocation of limited resources. The project was launched in 2018; however, the strategy was significantly revised in 2021 to enhance its operational effectiveness. The project’s strategy aligns with recommendations from the Global Measles and Rubella Strategic Framework 2021-2030, which emphasize improved use of surveillance data to guide intervention decisions and prioritizing vaccination efforts in geographic areas with the most significant immunity gaps [13].

In this work, we aim to demonstrate the value and effectiveness of targeted preventive activities, examine strategies for prioritizing actions in the face of numerous alerts, and assess the overall impact of interventions on measles epidemics. Specifically, we evaluate both the performance of the epidemiological tools used to guide operational decision-making during 2021-2023 and the outcomes of the preventive and reactive vaccination efforts. The goal is to draw lessons that can strengthen future epidemic responses and provide insights that may be applicable in other geographic and epidemiological contexts.

## Material and Methods

### Overview of Urgepi activities

The Urgepi project operates across all 68 health zones in the provinces of Haut-Katanga, Haut-Lomami, Lualaba, and Tanganyika. To maximize the impact of the limited operational capacities, the project follows a risk-targeted approach by allocating more resources to health zones at higher risk of large measles outbreaks - designated as “priority health zones.”

The prioritization strategy guides several components of the project (Figure 1). Preventive vaccination campaigns are conducted in selected priority health zones. In addition, Urgepi reinforces surveillance and applies more sensitive alert thresholds in priority health zones to enable earlier detection of measles epidemics and facilitate timely response.

**Figure 1.**
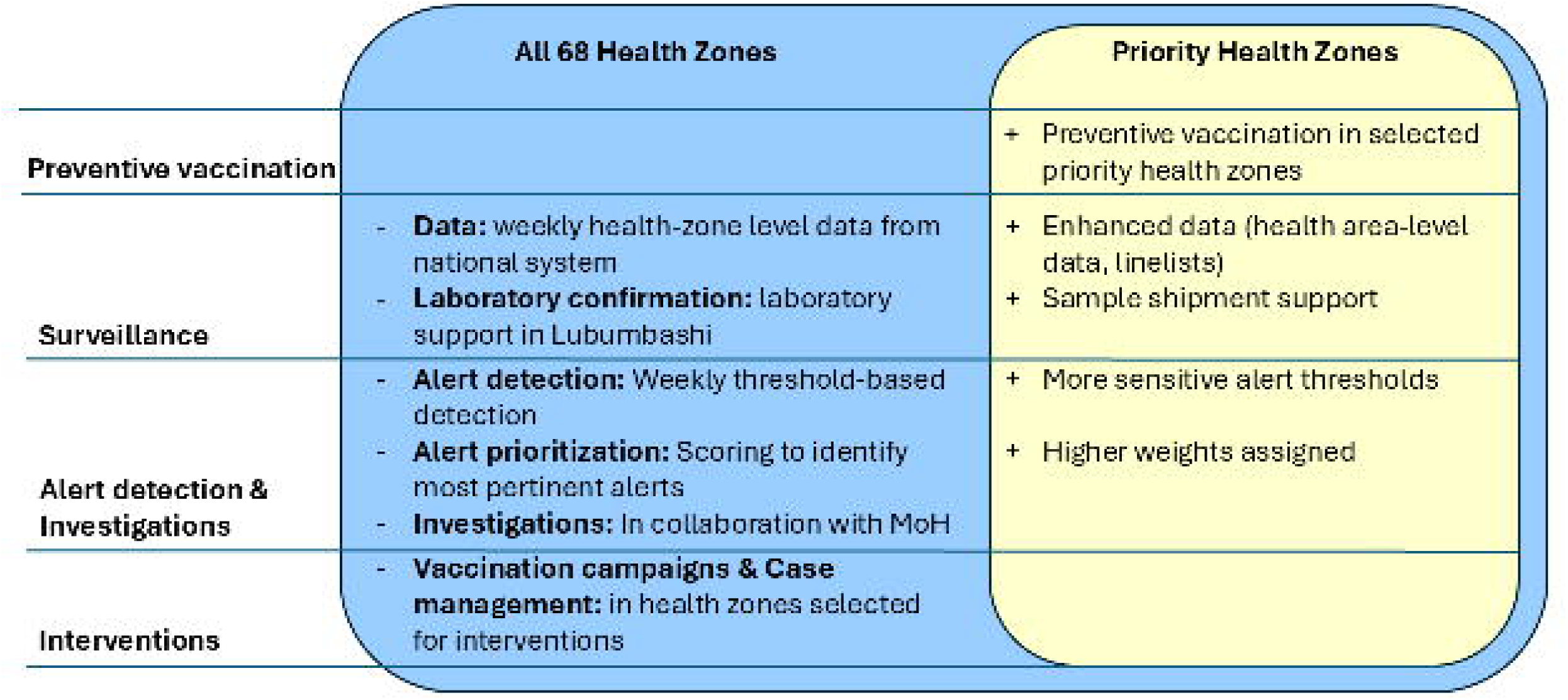
Risk – targeted strategy of the Urgepi project. Comparison of routine versus enhanced activities across health zones.

During peak periods of regional measles transmission, when the number of alerts can exceed operational capacity, an alert prioritization algorithm is applied to maximize the impact of the outbreak response. This algorithm assigns a quantitative score to each zone in the alert stage. Health zones are subsequently selected for field investigation and response based on the alert prioritization scores and operational discussions. Once an alert is selected, a field investigation team is deployed to confirm the magnitude / severity of the epidemic, better understand the local context, and plan an appropriate intervention, typically involving reactive vaccination and support for case management.

### Selection of “priority health zones”

The selection of priority health zones evolved over the course of the project from 2021 to 2023. Initially, in 2021, selection was based on a consensus rank, supplemented by local risk assessments and consideration of the project’s operational constraints. In 2022, the list was revised to ensure a more balanced distribution of priority health zones across all four provinces. Thereafter, the selection was updated every six months to reflect recent vaccination campaigns and completed epidemics. Health zones no longer requiring targeted support were replaced by others based on their risk. Further details on the selection process and evaluation of selection are provided in the Supplementary Material (Supplementary Table S1, Supplementary Figure S1, Supplementary Figure S2, Supplementary Figure S3 and Supplementary Figure S4).

### Surveillance

The Urgepi project includes a surveillance component to monitor the measles situation across all 68 health zones. Surveillance is conducted in collaboration with the Direction of Epidemiological Surveillance of the Ministry of Health (MoH) and relies on data from the national surveillance system. The Division Provinciale de la Santé (DPS) shares weekly reports of suspected measles cases and deaths by health zone with the Urgepi project. Additionally, the project receives weekly laboratory confirmation data from the Grand Laboratoire de Lubumbashi.

In priority health zones, the project strengthens surveillance by providing training on surveillance procedures, technical and, when needed, financial support for epidemic investigations, and logistical and financial assistance for laboratory confirmation of measles cases. The Urgepi project maintains regular direct contact with local authorities in these priority zones, who share detailed information on the distribution of suspected cases at the health area level (admin level 3), as well as linelist data when available.

### Alert thresholds and alert prioritization

The Urgepi project uses two alert definitions based on the health zone’s priority status. In non-priority health zones, an alert is triggered by any of the following: 20 suspected measles cases in one week, 35 suspected cases over the past three weeks, or 3 confirmed measles cases over the past three weeks. In priority health zones, the criteria are more sensitive: 10 suspected cases in one week, 15 suspected cases over three weeks, or 1 confirmed case. These alert definitions were developed based on an evaluation of historical surveillance data, aiming to strike a balance between timely detection of measles epidemics and limiting false alerts that could overwhelm the project’s operational capacity.

The alert definitions were selected to align with the Urgepi project’s resources and operational capacities and therefore differ from the alert thresholds used by the MoH. Both approaches are complementary: The MoH applies more sensitive thresholds, triggering immediate investigations into small clusters of suspected cases given its presence across all health zones. In contrast, Urgepi investigations are more resource-intensive, requiring additional personnel and transportation to affected areas. Accordingly, Urgepi uses less sensitive alert definitions that focus on signals with a higher likelihood of indicating an emerging epidemic.

Although all health zones in alert stage warrant attention, prioritization of alerts may be necessary when these outnumber the operational capacity. Prioritization helps determine the allocation of response efforts, ensuring that interventions first target areas where action is expected to prevent the greatest number of cases and related deaths. The Urgepi project therefore applies an alert prioritization algorithm to each health zone in alert stage. This algorithm incorporates both epidemiological indicators - such as the number of suspected cases over the past three weeks, trend in case numbers, case fatality rate, and laboratory confirmation of an outbreak - and contextual factors, including whether the health zone is a priority health zone, the timing and scale of the last epidemic, routine vaccination coverage, and the epidemic status of neighboring health zones. A detailed description of the alert scoring algorithm is available in Supplementary material Table S2. Alert scores may range from 0 to 16, with 16 indicating the highest priority. These scores are reviewed weekly to support operational decision-making.

### Preventive vaccination and epidemic response activities

During periods without widespread regional epidemics as during the year after national supplementary immunization activities, the Urgepi project, in collaboration with the MoH, carries out preventive measles vaccination campaigns in selected priority health zones. In 2021, the target age group for vaccination varied based on discussions with local health authorities and included all children aged 6 to 59 months (in Dilolo, Kasaji, Kinkondja, Mukanga, and Kabongo health zones) or 6 to 23 months (in Manono and Kiyambi), or selectively targeted unvaccinated children aged 9 to 23 months (in Pweto and Kilwa).

In the event of an ongoing measles epidemic, the Urgepi project implements a comprehensive response that includes free case management for measles patients, including hospitalized pediatric patients, and a reactive vaccination campaign. These vaccination campaigns are conducted in collaboration with the MoH and target all children within the most affected age group, typically those aged 6–59 months. A health zone becomes eligible for a reactive vaccination campaign once the epidemic is officially declared by the MoH, which generally follows laboratory confirmation of at least three measles cases.

### Data analysis

#### Describing measles epidemics and interventions

Measles epidemics were described based on the national surveillance data (IDSR) [3] for the four provinces Haut Katanga, Haut Lomami, Lualaba and Tanganyika during 2021-2023. MSF interventions (preventive and reactive) were described during the same period based on the intervention database of the Urgepi project.

#### Evaluating the impact of measles preventive vaccination

We compared the number and proportion of health zones that experienced large measles epidemics (defined as ≥500 suspected cases), as well as median attack rates, between health zones that received preventive vaccination in 2021 (n = 9) and those that were prioritized in 2021 but did not receive either preventive or early reactive vaccination (n = 10). Two priority health zones (Mitwaba and Mufunga Sampwe) were excluded from the analysis, as they received early reactive vaccination campaigns led by the MoH before many cases was reported. We estimated the statistical significance of the difference in proportions using Fisher’s exact test, and for the difference in median attack rates using Wilcoxon rank-sum test.

For health zones that experienced large epidemics despite preventive vaccination, we described the spatial distribution of suspected cases by health area (admin-3 level).

#### Evaluating alert prioritization for measles epidemic response

To evaluate the performance of the alert prioritization algorithm, we calculated weekly alert scores for each health zone in the alert stage during 2021–2023.

To align the timescale across epidemics, we identified the first week with ≥20 suspected measles cases in each health zone as the standardized starting point of the epidemic. The identification of the first ≥20-case week was initially done automatically and then manually reviewed. If the identified week was clearly disconnected from the main epidemic curve, we used the second week with ≥20 cases as the epidemic start instead. We then extracted the maximum alert score from the period before the epidemic up to eight weeks after this initial threshold week. For health zones in alert that never reached the ≥20 suspected cases per week threshold during the study period, we extracted the maximum alert score recorded at any point between 2021 and 2023.

We calculated the median epidemic attack rate and interquartile range (IQR) for each alert score and assessed statistical significance using the Kruskal-Wallis test. We further estimated the probability of experiencing a large epidemic (≥500 suspected cases within the 2021-2023 time window) by alert score using a generalized linear model (GLM) with a basic spline to account for non-linear effects and estimated the p-value of association using a likelihood ratio test.

Health zones with successful preventive vaccination or early reactive vaccination (i.e. health zones with preventive or reactive vaccination campaigns that never reached 500 cases) were excluded from the analyses. Health zones that had not experienced alerts were also excluded automatically, as for these no alert score was estimated.

We repeated the analysis considering alert scores only up to 4 weeks after the weekly threshold of 20 cases was first reached (Supplementary material).

#### Evaluating the timing and impact of reactive vaccination campaigns

To evaluate the timing of reactive vaccination campaigns, we quantified delays between the first week of ≥20 notified suspected cases and the start of vaccination activities.

We compared the median and IQR of the number of cases, AR, and epidemic duration between health zones that implemented Urgepi-supported reactive vaccination campaigns and those that reported ≥20 suspected cases in at least one week but did not conduct a reactive vaccination campaign. Epidemic duration was defined as the time between the first and last week with ≥20 reported cases. As with the first ≥20-case week, the last such week during 2021–2023 was initially identified automatically and then manually reviewed and adjusted if it appeared disconnected from the main epidemic curve. We tested statistical significance of differences using the Wilcoxon rank-sum test.

In addition, we explored the correlation between the delay in vaccination campaign initiation and the case numbers or AR observed in each health zone using a generalized linear model (GLM) with negative binomial link for cases and a linear regression for AR.

### Patient and public involvement

Patients and/or the public were not involved in the design, conduct, reporting, or dissemination plans of this research.

## Results

### Measles epidemics and MSF interventions in the provinces of Haut-Katanga, Haut-Lomami, Lualaba and Tanganyika during 2021-2023

During 2021-2023, a total of 67 595 suspected measles cases, including 681 suspected measles deaths, were notified in the four provinces Haut Katanga, Haut Lomami, Lualaba, and Tanganyika (Figure 2 A). The most affected province was Tanganyika with 20 226 suspected cases and an AR of 583 per 100 000 population, followed by Haut Lomami, Lualaba, and Haut-Katanga (Table 1). While all 68 health zones notified at least one suspected measles case, large epidemics with ≥500 suspected cases were observed in 54% (37/68) of the health zones (Figure 2 B).

**Figure 2.**
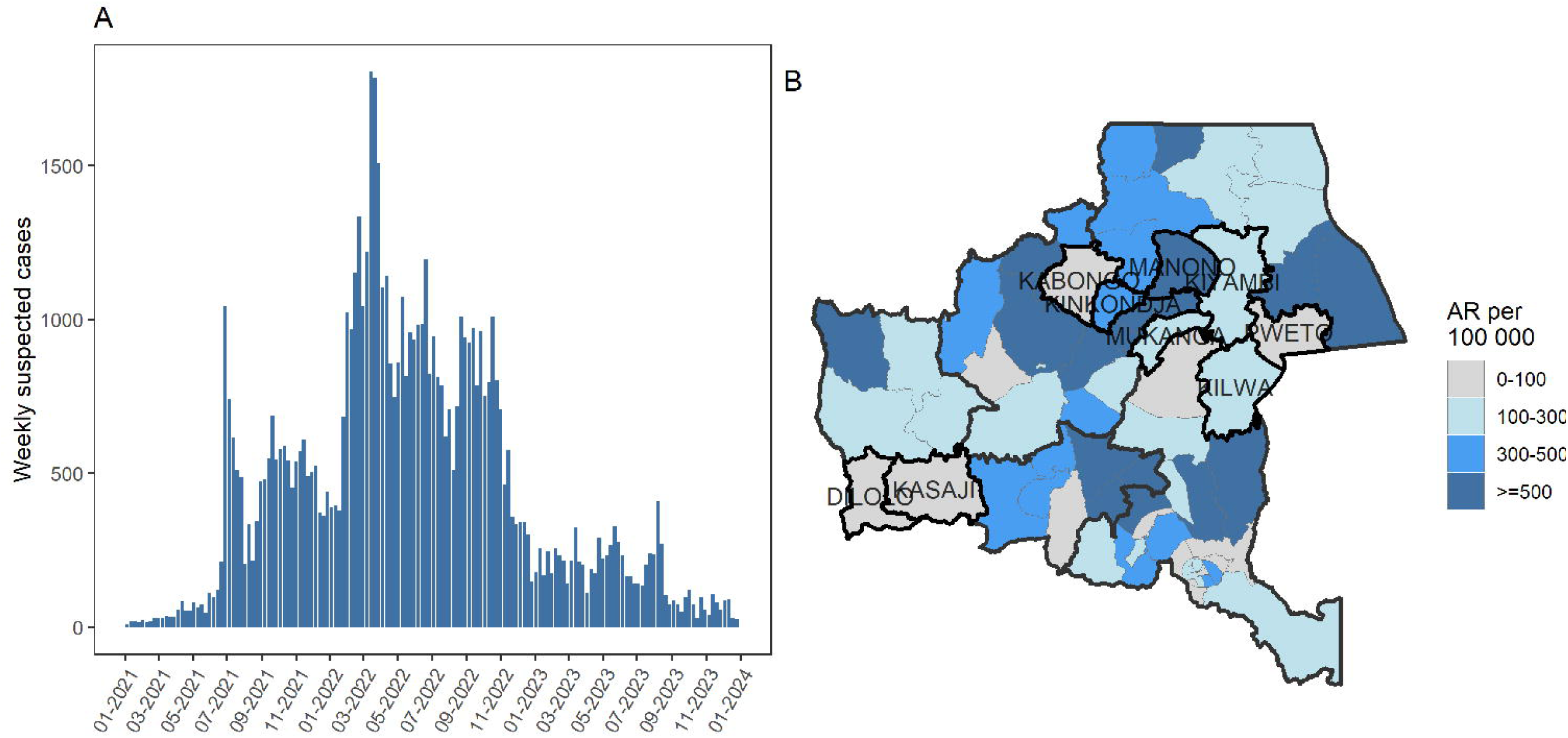
Suspected measles cases notified by week (A) and by health zone (B) during 2021-2023. The 9 health zones included in preventive vaccination activities are indicated by their name.

**Table 1.**
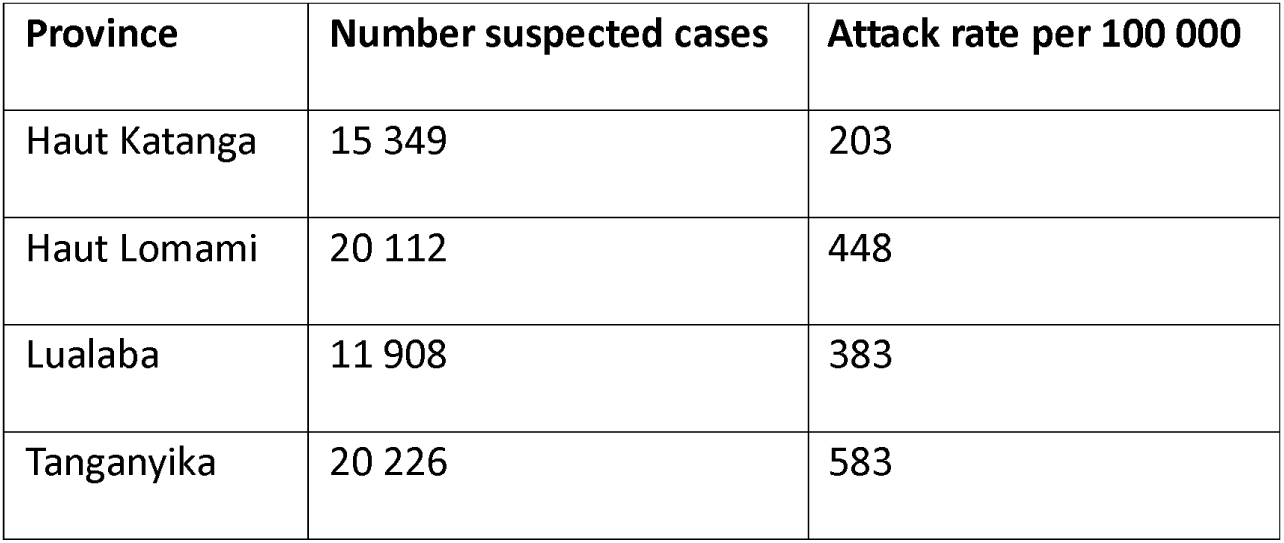
Number of suspected measles cases and attack rate per 100 000 during 2021-2023 by province.

| Province | Number suspected cases | Attack rate per 100 000 |
| --- | --- | --- |
| Haut Katanga | 15 349 | 203 |
| Haut Lomami | 20 112 | 448 |
| Lualaba | 11 908 | 383 |
| Tanganyika | 20 226 | 583 |

During June-October 2021, the Urgepi project in collaboration with the MoH implemented preventive measles vaccination campaigns in 9 health zones (Figure 2). These vaccination activities took place before the occurrence of epidemics in the respective health zones. Over 300 000 children were vaccinated during these campaigns, with administrative vaccination coverage ranging from 100 to 110%.

Between June 2021 and September 2023, the Urgepi project responded to 24 measles epidemics, implementing reactive vaccination campaigns and case management activities in collaboration with the MoH. Five interventions took place in 2021, 16 in 2022 and 3 in 2023. More than 1 million children aged 6-59 months were vaccinated (administrative coverage of 93-118%), and 22 626 measles patients (including 3 280 severe measles patients) were treated in the supported health facilities.

In addition, the MoH implemented reactive vaccination campaigns in 20 health zones.

### Evaluating the impact of preventive vaccination campaigns

While 90% (9 out of 10) of unvaccinated priority health zones experienced large measles epidemics (≥500 cases) (Supplementary material Figure S5), only 33% (3 out of 9) of vaccinated zones reported this level of cases (p= 0.02) (Supplementary material Figure S6). This suggests that preventive vaccination resulted in a 3 times lower risk of outbreaks, and likely averted large epidemics in 5 high-risk health zones. The median attack rate in the 10 unvaccinated health zones was 417 per 100,000 population (IQR 306-514 per 100 000), compared to 127 per 100,000 in the vaccinated high-risk health zones (p=0.02) (IQR 34 – 254 per 100 000).

In the 3 high-risk health zones that experienced large epidemics despite preventive vaccination, the spatial distribution of measles cases ranged from geographic pockets (Kilwa health zone) to a wide scale epidemic (Manono health zone) (Supplementary material Figure S7).

### Evaluating the prioritization of alerts for an MSF-supported measles response

During 2021-2023, alerts were detected in 66 /68 health zones. Up to 8 weeks after the first 20 cases were reported, maximum alert scores across health zones ranged from 6 to 16, with a median score of 12 (IQR 9 -13). The probability of large measles epidemics increased significantly from 3.7% (95%CI 0.0-75.4) for alerts with a score of 6 to 96.3% (95%CI 24.3-100.0) for alerts with a score of 16 (p=0.002) (Figure 3). An increase in median attack rates was also observed, however was not statistically significant (p=0.10) (Figure 3). Among the 24 health zones where MSF responded to measles epidemics, maximum alert scores ranged from 8 to 16.

**Figure 3.**
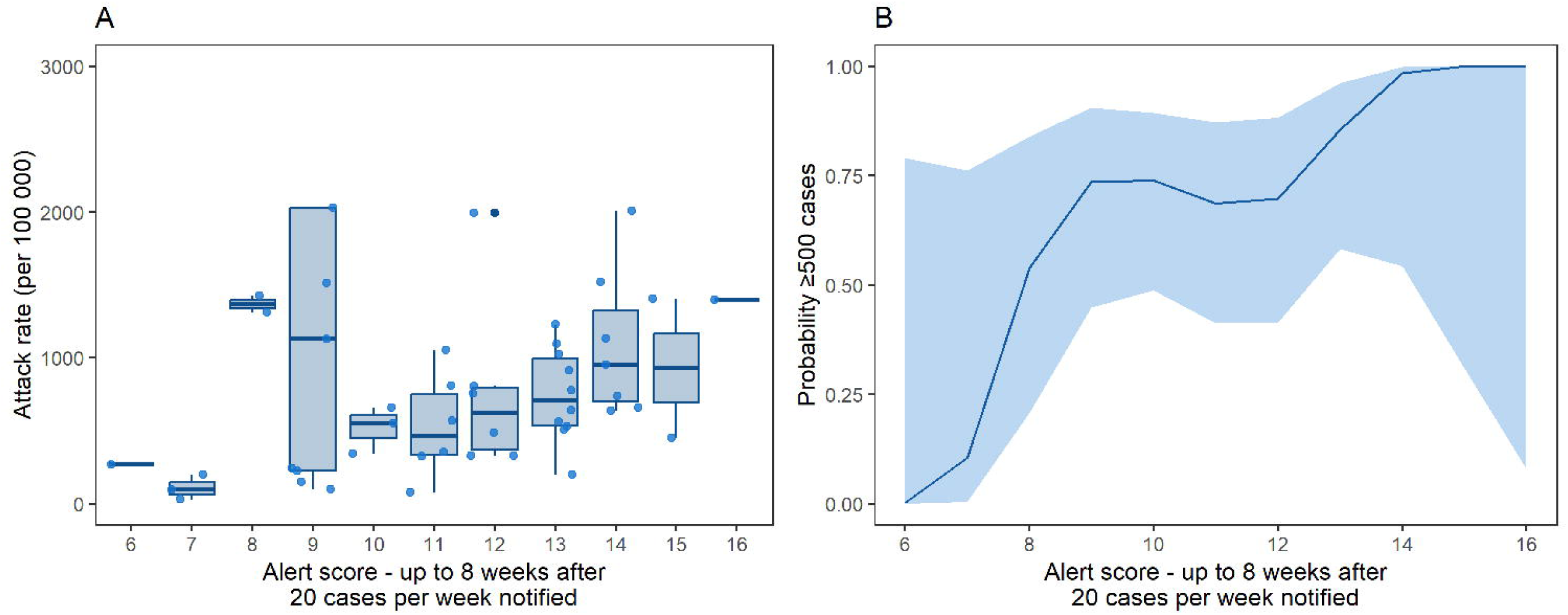
Attack rate per 100 000 (A) and probability of large epidemics (≥500 cases) (B) by alert score up to 8 weeks after the weekly threshold of 20 cases was first reached. For health zones without notification of ≥20 cases per week the maximum score was considered.

The increase in attack rates and probability of large epidemics was less pronounced when considering alert scores only up to up to 4 weeks after the first 20 cases were reported (Supplementary material Figure S8).

### Evaluating the timing and impact of reactive vaccination campaigns

Most reactive vaccination campaigns (16/24; 66.7%) were implemented in priority health zones and campaigns were launched a median of 7 weeks after the weekly threshold of 20 cases was first reached (IQR: 5-10 weeks; range: 1-14 weeks).

In health zones where Urgepi responded to measles epidemics, the number of reported cases ranged from 182 to 5 151, with ARs between 110 and 1 432 per 100 000 population. Delays in launching reactive vaccination campaigns were associated with higher case counts (p=0.001) and attack rates (p=0.001). While the estimated mean number of cases increased from 710 cases (95%CI 470-1 071) for health zones with 14 days vaccination delay to 3 126 cases (95%CI 2 025-4 827) for health zones with 90 days delay, the AR increased from 374 per 100 000 (95%CI 176 -572 per 100 000) to 1 002 per 100 000 population (95%CI 793 – 1211 per 100 000) (Supplementary material Figure S9).

To evaluate the impact of reactive vaccination activities, we compared the number of cases and attack rates in 24 health zones with Urgepi reactive vaccination campaigns with 16 health zones that reported ≥20 cases in at least one week but did not conduct health zone wide reactive vaccination campaigns. Surprisingly, both the median number of cases and attack rates were higher in the health zones with Urgepi response. Vaccinated health zones: median of 1 393 cases (IQR: 888–1 908); median attack rate of 700 per 100,000 (IQR: 440–850); non-vaccinated health zones: median of 400 cases (IQR: 98–857); median attack rate of 141 per 100,000 (IQR: 49–303) (Figure 4). The differences in case numbers and attack rates were both statistically significant (p<0.001). However, these estimates may be biased, as the Urgepi interventions also included free case management, support for patient referrals, and enhanced surveillance, which likely increased case detection and reporting.

**Figure 4.**
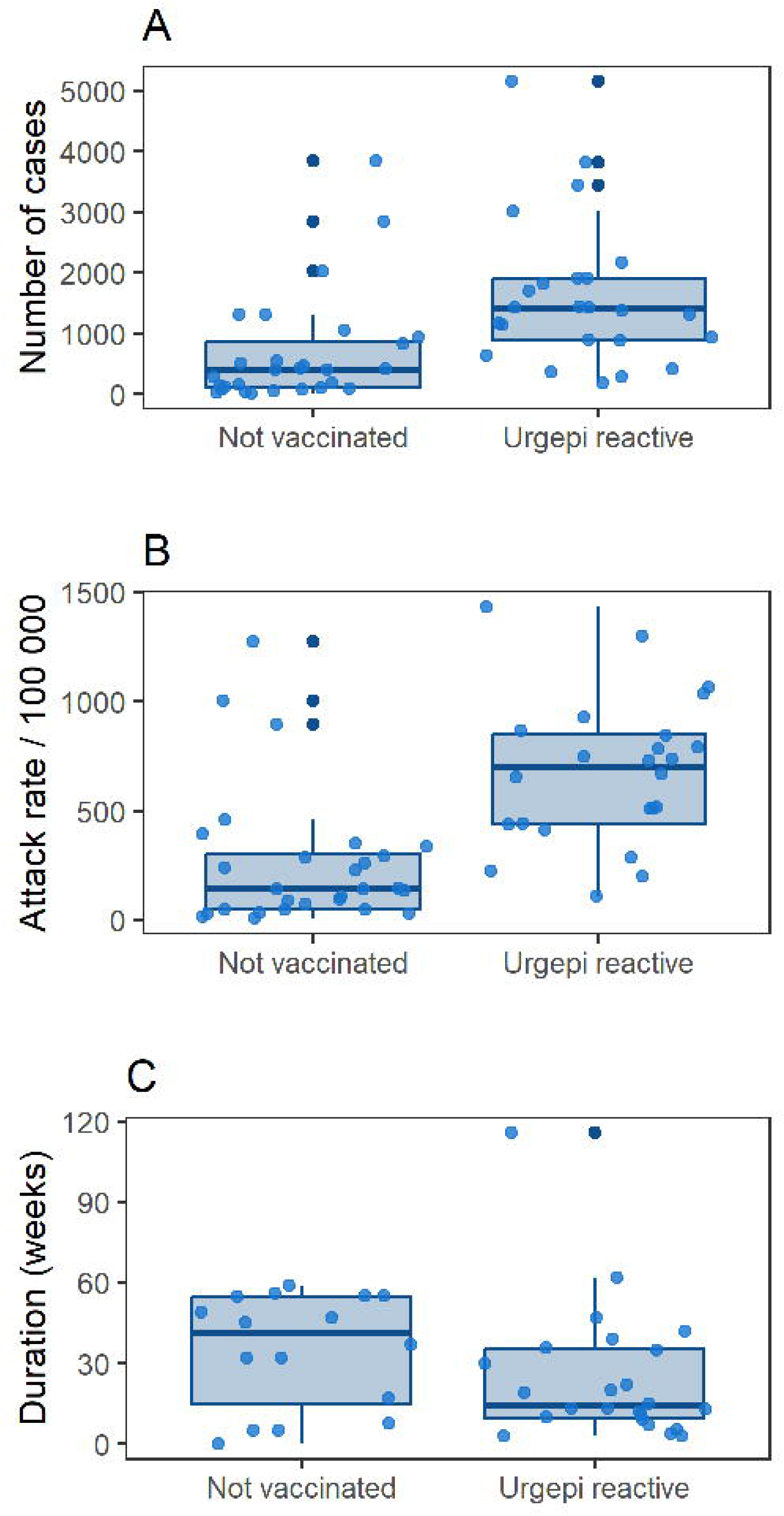
Number of cases (A), attack rates per 100 000 population (B), and duration of epidemics (C) in 24 health zones with Urgepi reactive vaccination campaigns, compared to 16 health zones that reported ≥20 cases in at least one week but did not implement reactive vaccination campaigns.

We also compared the duration of measles epidemics, defined as the time between the first and last weeks with at least 20 suspected cases. Epidemics in the 24 health zones that implemented reactive vaccination campaigns had a shorter median duration of 14 weeks (IQR: 10–35 weeks), compared to 41 weeks (IQR: 15–55 weeks) in the 16 zones without such campaigns, although this tendency was not statistically significant (p=0.10) (Figure 4).

## Discussion

Reactivity is key to reducing the impact of an epidemic. Detecting alerts early, while keeping the risk of false alerts low, is essential for mounting an effective and timely response. However, when multiple epidemics occur simultaneously and resources are limited, prioritization becomes equally important to ensure the most efficient use of available capacities. In this context, the risk-targeted Urgepi approach, which strategically combined preventive and reactive interventions, played a key role in averting or reducing the scale of measles epidemics across several health zones in the Katanga region between 2021 and 2023. The evaluation and capitalization of the project’s activities demonstrated the effectiveness of these strategies but also identified areas for further improvement.

Preventive vaccination in identified high-risk health zones is a powerful strategy, especially in hard-to-reach areas where timely response during reactive campaigns can be particularly challenging or even impossible during the rainy season. It is also particularly valuable in areas where resistance to vaccination is high, as it allows more time to identify these challenges early and engage with communities before an outbreak occurs. The preventive campaigns carried out in 2021 had a notable impact: six of the nine vaccinated health zones remained free of epidemics during the epidemic period, despite being surrounded by affected areas. In contrast, nearly all non-vaccinated priority zones experienced epidemics.

Despite being targeted for vaccination, three health zones still experienced measles epidemics, highlighting persistent gaps in vaccination coverage. These situations point to the need for better identification and understanding of the barriers to achieving high coverage and the adoption of more locally adapted strategies. The case of Manono health zone, where more than 5000 cases were reported across all health areas despite Urgepi supported preventive and reactive vaccination campaigns, illustrates these challenges clearly and underscores the importance of better preparation and deeper contextual analysis to guide future interventions. Such insights can be gained through qualitative or mixed methods studies studies focusing on vaccination barriers, including community perceptions, access challenges, and operational constraints [14–17]. Indeed, a qualitative study conducted in 2023 in Manono revealed that both distrust in the vaccine itself and skepticism toward the institutions involved in vaccination contributed to vaccine refusals and low coverage [18]. Additionally, a vaccination coverage survey conducted six months after the preventive campaign—and one month after a reactive campaign in 2022—found that less than 60% of targeted children were reached during both campaigns. The main reasons for missed vaccinations were parents’ unavailability during the campaign period and vaccine refusal [19].

For reactive vaccination activities, the alert scoring algorithm proved valuable in prioritizing epidemics for intervention, with higher alert scores correlating to larger epidemic sizes. This approach effectively guided decision-making, demonstrating that alert scoring can support more efficient, targeted responses. In prioritizing ongoing epidemics, we aimed to use straightforward methods that avoid complex computational algorithms, making them suitable for resource-limited settings. A previous literature review highlighted the scarcity of simple and dynamic prioritization tools to guide epidemic responses amid evolving epidemiological situations [20]. Here, we illustrate how such practical approaches can enhance timely and informed decision-making in outbreak response.

We demonstrated that Urgepi reactive vaccination was correlated with a reduction in the duration of epidemics, but not with lower epidemic size. When evaluating the impact of interventions, epidemic size alone is not a reliable indicator. The risk-targeted approach, combined with free case management, may lead to higher reported case numbers and attack rates in health zones where MSF conducted reactive vaccination campaigns, compared to those without such interventions. However, a clear impact of our interventions was observed on the duration of epidemics: the median duration was reduced by more than 50% compared to epidemics without interventions. This may correspond to a median reduction of cases by 50%, given the generally symmetrical shape of epidemic curves. As expected, interventions implemented with shorter delays had a greater impact, underscoring that to maximize effectiveness, delays in launching reactive campaigns must be shortened [21].

In resource-limited settings, the Urgepi project’s risk-targeted approach proved valuable in improving measles prevention and control. Preventive campaigns helped avert epidemics in several high-risk zones, while reactive responses, guided by a simple alert scoring tool, contributed to shorter epidemic durations. The findings highlight the importance of timely interventions, context-specific strategies, and deeper understanding of local barriers to vaccination to maximize the effectiveness of outbreak response.

## Data Availability

The Medecins Sans Frontieres (MSF) operational data underlying this study are available on request, in accordance with the legal framework set forth by MSF data sharing policy (Karunakara U, PLoS Med 2013). MSF is committed to share and disseminate health data from its programs and research in an open, timely, and transparent manner in order to promote health benefits for populations while respecting ethical and legal obligations towards patients, research participants, and their communities. The MSF data sharing policy ensures that data will be available upon request to interested researchers while addressing all security, legal, and ethical concerns. All readers may contact to request data. The underlying surveillance data are available upon reasonable request from the Epidemiological Surveillance Directorate, Measles-Rubella Data Analysis Unit, Kinshasa, CD

## Acknowledgements

We would like to thank all intervention and research teams of the MoH DRC and MSF involved in surveillance activities, preventive vaccination, and outbreak response during the study period.

## Ethics statement

This research fulfilled the exemption criteria set by the Médecins Sans Frontières Ethics Review Board for a posteriori analyses of routinely-collected clinical data and thus did not require MSF ERB review. It was conducted with permission from Medical Director, Operational Center Paris, Médecins Sans Frontières.

## Contribution statement

BN, CE, MJF, LS, EG, CM, KP and IAQ conceptualized and designed the study. HS, CM, GM, NM, DM, JMK, PBM, FK, WLN, JM, SA, EPS, VN, VD, RAA, AC, NL coordinated and implemented study and routine surveillance activities. BN and CE wrote the statistical analysis plan. BN, HS, DM cleaned and analyzed the data. BN, CD, CM, and IAQ drafted the manuscript with substantial input from all authors. All authors reviewed the manuscript and were responsible for the decision to submit. BN is the guarantor. Microsoft 365 Copilot was used for copyediting and language refinement during manuscript preparation. All AI outputs were reviewed and approved by the authors before inclusion.

