## Supplementary material for "Risk-targeted interventions for measles control-Lessons from an emergency response project in the Katanga Region, Democratic Republic of the Congo (2021-2023)"

1. **Identification of high-risk health zones**
   1.1. **Prioritization methods**

In the beginning of 2021, 21 health zones were selected based on a consensus rank of three indicators [Ferrari M, unpublished]:

1. historical vaccination coverage (a population weighted estimate based on the 2013-14 Demographic Health Survey) [1]

2. historical number of unvaccinated children - this is population under 5 years old * (1- historical vaccination coverage under 5 years)

3. estimated number of unvaccinated children in 2021 (combining the historical coverage with campaigns that have occurred since the DHS survey)

The ranks of health zones were then summed up for each of these indicators (e.g. a health zone that is ranked as highest priority of each the indicators would receive a score of 3).

The selection from 2021 focused mainly on the province of Haut Katanga due to logistical reasons (Table S1). In the beginning of 2022, 20 health zones were selected that were for collaborative reasons equally distributed among the four provinces. The selection also followed the risk model ranking for each province, excluding however those health zones that had already benefitted from preventive or reactive vaccination activities among children 9-59 months, as well as those health zones that already experienced an epidemic in 2021. Additional logistical aspects (e.g., accessibility) were considered in the final selection. The list was then revised in mid-2022 following the same logic, retaining 9 health zones. After the initial selection of 21 health zones, by mid-2022 a total of 33 health zones and by end of 2023 a total of 37 health zones had been considered as priority health zones during the 3-year period.

**Table S1. Urgepi priority health zones.**

| Period | Nb Priority health zones | Priority health zones |
| --- | --- | --- |
| January-December 2021 | 21 | **Haut Katanga:** Kampemba, Kilela Balanda, Mitwaba, Pweto, Kambove, Kapolowe, Kasenga, Kilwa, Lukafu, Mufunga Sampwe, Sakania  **Haut-Lomami:** Kabongo, Mukanga, Kinkondja, Bukama  **Lualaba:** Kasaji, Dilolo, Manika  **Tanganyika:** Kiyambi, Manono, Kalemie |
| January- June 2022 | 20 | **Haut-Katanga:** Pweto, Kilwa, Kasenga, Mitwaba, Kipushi  **Haut-Lomami:** Kaniama, Kayamba, Mulongo, Bukama, Lwamba  **Lualaba:** Sandoa, Kalamba, Mutshatsha, Kapanga, Kafakumba  **Tanganyika:** Kalemie, Kiyambi, Manono, Ankoro, Moba |
| July - December 2023 | 9 | **Haut-Katanga:** Kafubu,  **Haut-Lomami:** Kayamba, Songa, Kinda, **Lualaba:** Sandoa, Kalamba, Kapanga, Kafakumba, Lubudi |

1.2. **Evaluating the prioritization of health zones for preventive measles activities**

**Methods- data analysis**

To retrospectively evaluate the selection of priority health zones, we assessed the proportion of large measles epidemics (≥500 cases) among health zones that were prioritized (i) up to the end of 2021, (ii) mid-2022, and (iii) the end of 2023, and compared it to the proportion of large epidemics among health zones that had not yet been prioritized at that time. We used cumulative prioritization periods because health zones were removed from the priority list once they either experienced an epidemic or benefited from a large-scale vaccination campaign. Similarly, we compared the median number of cases and the median attack rate among priority health zones to non-priority health zones.

To identify a simpler prioritization method, we also tested among others an alternative method based on historical measles vaccination coverage among children <59 months old. The historical vaccination coverage by health zone was based on a Demographic health survey (DHS) in 2014 and derived from [1]. A Bayesian spatial model was used to estimate vaccination coverage at 1 km grid resolution. These estimates were combined with under-5-year-old population data to calculate the number of vaccinated children per grid cell. Health zone-level coverage was then derived by aggregating the number of vaccinated children and the total under-5 populations across all grid cells within each health zone. We considered a health zone as priority health zone if the historical vaccination coverage was ≤40%, which represents the lowest quartile of historical vaccination coverage in the Katanga region.

Excluded from the analyses were health zones with successful preventive vaccination or early reactive vaccination (i.e. health zones with preventive or reactive vaccination campaigns that never reached 500 cases).

**Results**

Among health zones prioritized during 2021, excluding those that benefited from successful preventive or early reactive vaccination, 92.3% (12/13) of health zones experienced a large epidemic (≥500 cases), compared to 64.1% (25/39) of health zones that had not yet been prioritized (Figure S1). Among health zones prioritized at any moment during 2021-2023, 84.0% (21/25) experienced a large epidemic, compared to 59.3% (16/27) of health zones that had never been prioritized (Figure S1). For all prioritization periods, the risk of large epidemics was higher among priority health zones than health zones that had not yet been prioritized. Similarly, the median number of cases and attack rates were higher among priority health zones than non-priority health zones (e.g. for the initial 2021 prioritization 1299 cases among priority health zones vs 833 cases among non-priority health zones) (Supplementary material Figure S2 and Figure S3).

To simplify the prioritization strategy for future activities of the Urgepi project, we also evaluated the prioritization of health zones based on low historical measles vaccination coverage (lowest quartile including health zones with ≤40% measles vaccination coverage, Supplementary material Figure S4). The proportion of theoretically prioritized health zones with large measles epidemics during 2021-2023 was slightly higher with 100% (14/14 health zones) than the actual prioritization and compares to a proportion of health zones with large epidemics of 60.5% (23/38 health zones) among theoretically non-prioritized health zones (Figure S1).

**Figure S1 Proportion of priority and non-priority health zones that experienced a large measles epidemic (≥500 cases) by prioritization period.** Additionally, we evaluated an alternative simplified prioritization based on low historical measles vaccination coverage (≤40%). This analysis is based on 52 health zones, excluded were 16 health zones that benefited from successful preventive or early reactive vaccination (through Urgepi or MoH) (i.e. vaccinated health zones that never reached ≥500 cases).

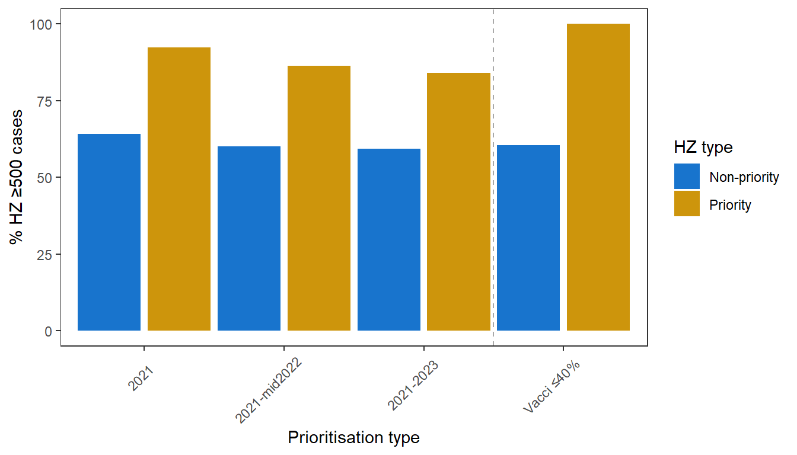

**Figure S2. Comparing the number of suspected measles cases (2021-2023) for health zones that were prioritized vs. not- yet prioritized during a certain period.** This analysis is based on 52 health zones, excluded were 16 health zones that benefited from successful preventive or early reactive vaccination (through Urgepi or MoH) (i.e. vaccinated health zones that never reached ≥500 cases).

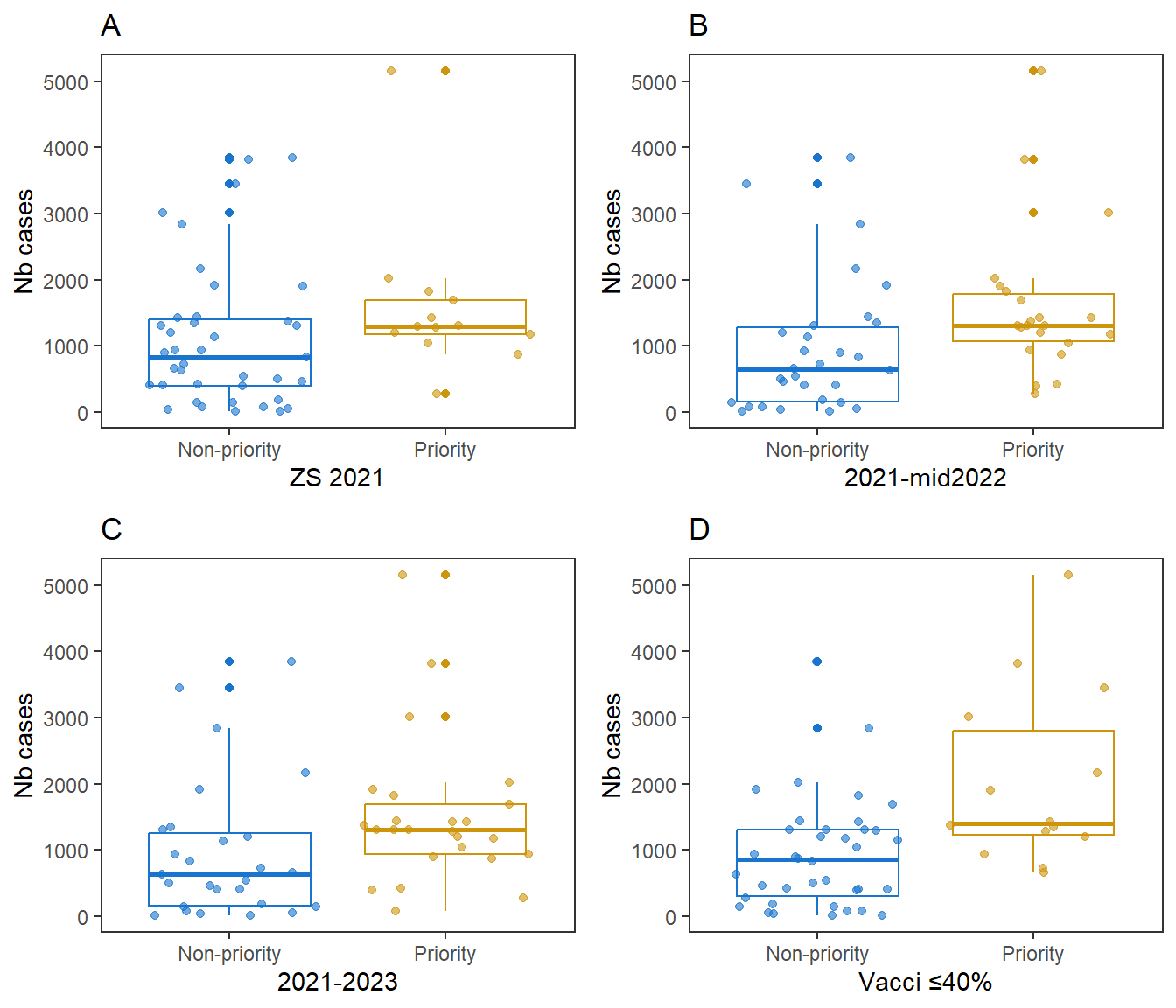

**Figure S3. Comparing the attack rates of measles (2021-2023) for health zones that were prioritized vs. not- yet prioritized during a certain period.** This analysis is based on 52 health zones, excluded were 16 health zones that benefited from successful preventive or early reactive vaccination (through Urgepi or MoH) (i.e. vaccinated health zones that never reached ≥500 cases).

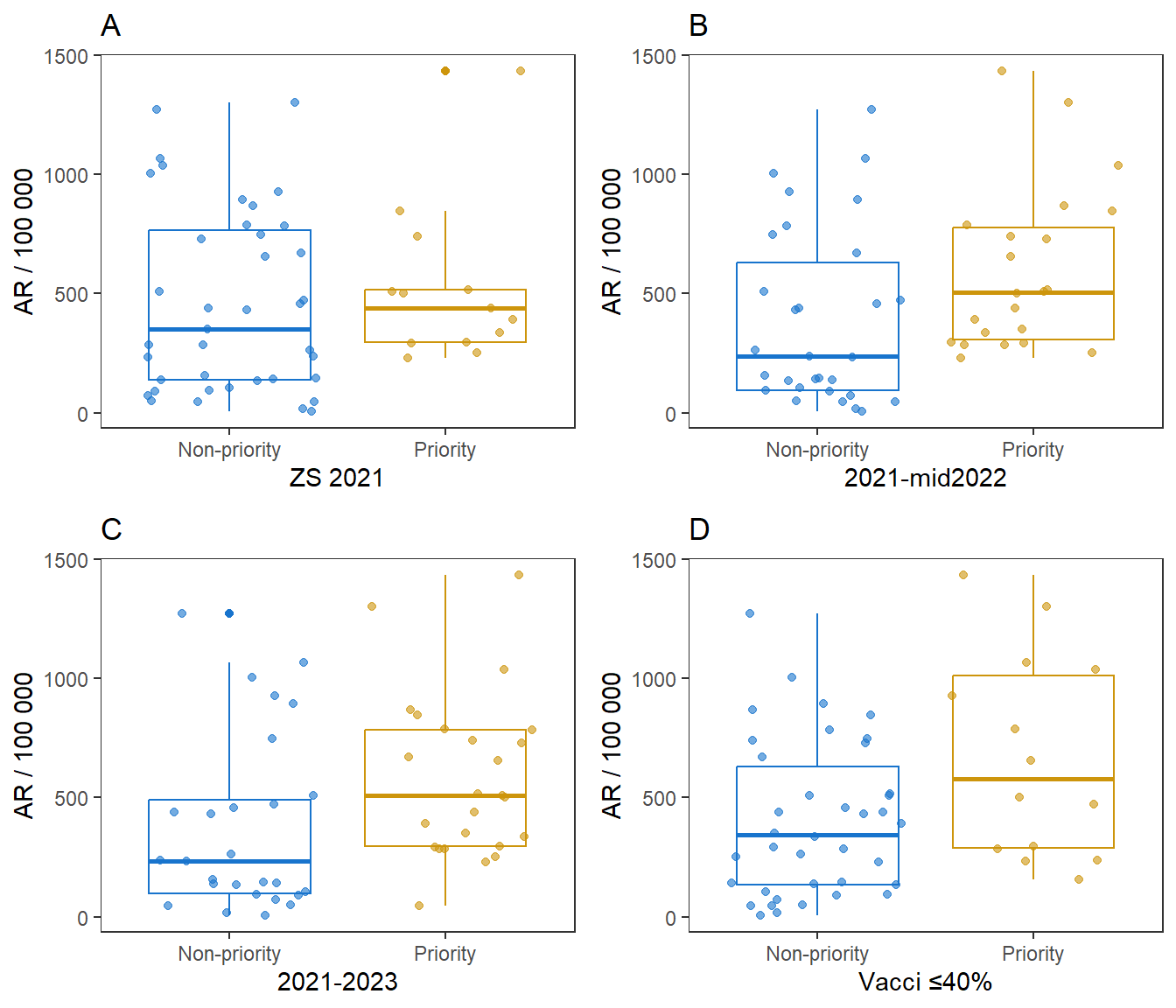

**Figure S4. Number of suspected measles cases (A) and AR per 100 000 (B) by quantile of historical measles coverage.** The first quantile includes heath zones with ≤40% vaccination coverage.

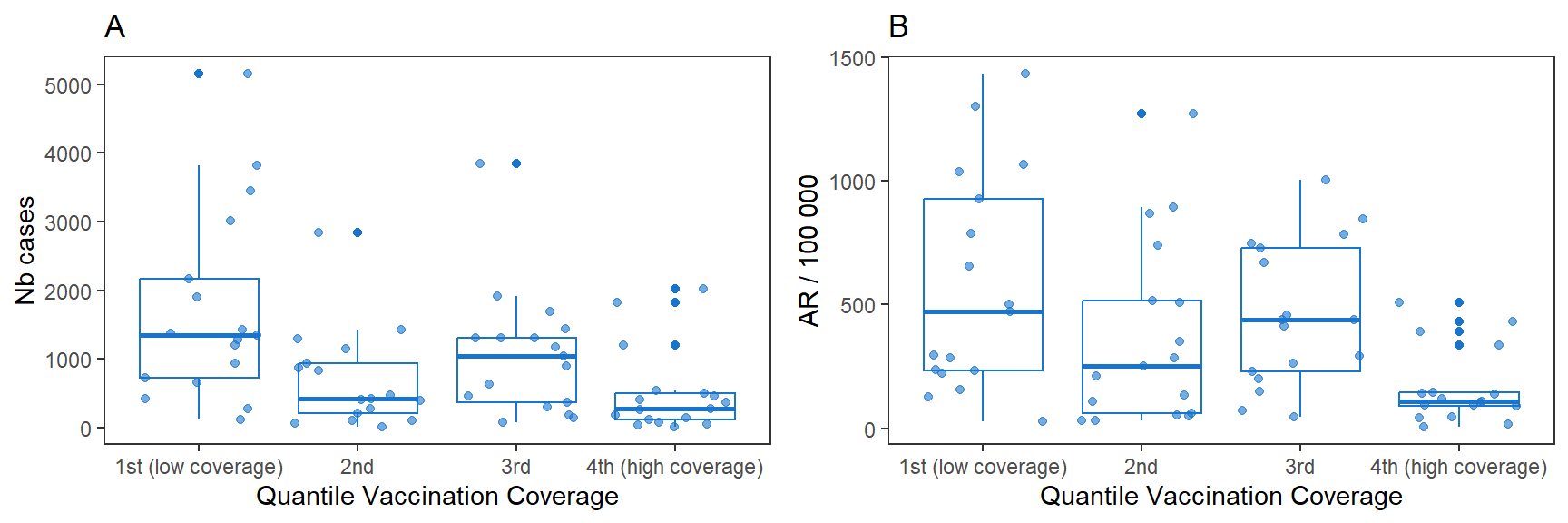

2. **Preventive vaccination – additional figures**

**Figure S5 Weekly reported suspected measles cases in health zones prioritized in 2021 but not targeted for preventive vaccination.** The health zones of Mitwaba and Mufunga Samwe were vaccinated by the MoH before a large epidemic was observed.

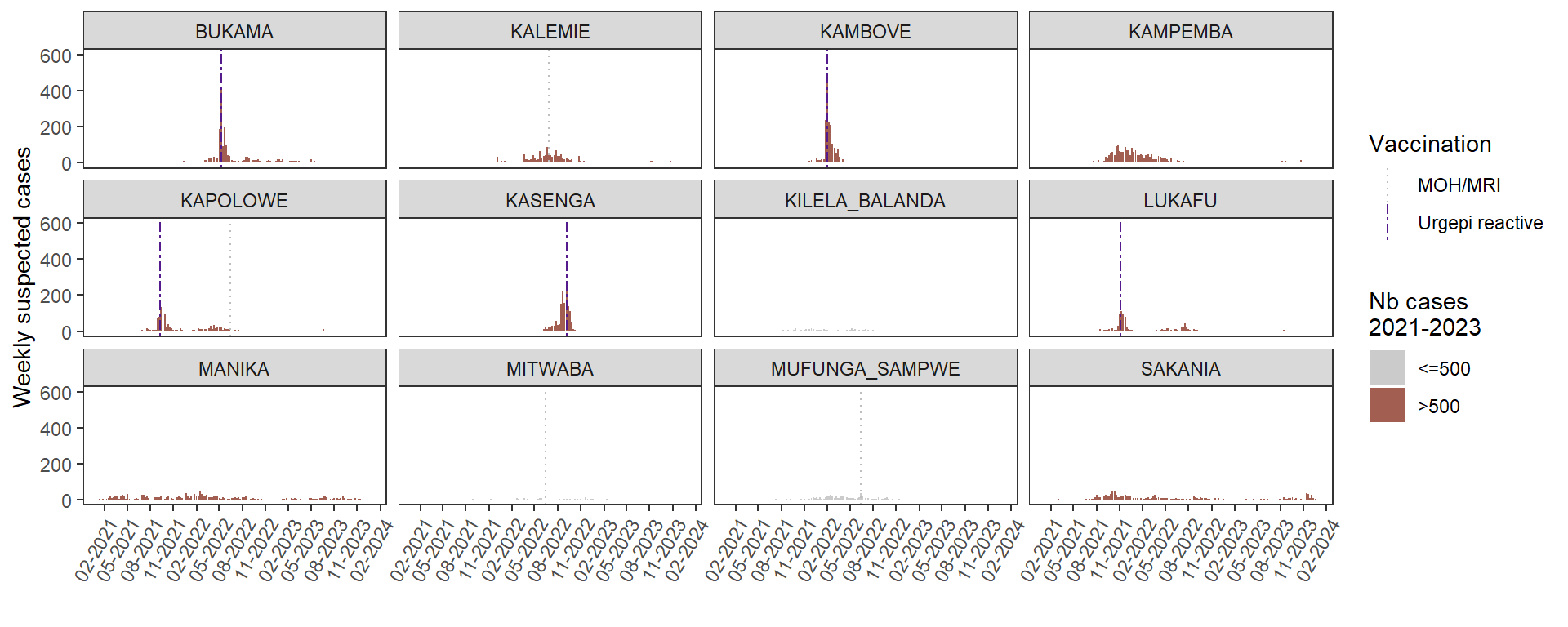

**Figure S6 Weekly reported suspected measles cases in health zones prioritized in 2021 and targeted for preventive vaccination.** (*vaccination 6-23 months, ** selective vaccination 9-23 months)

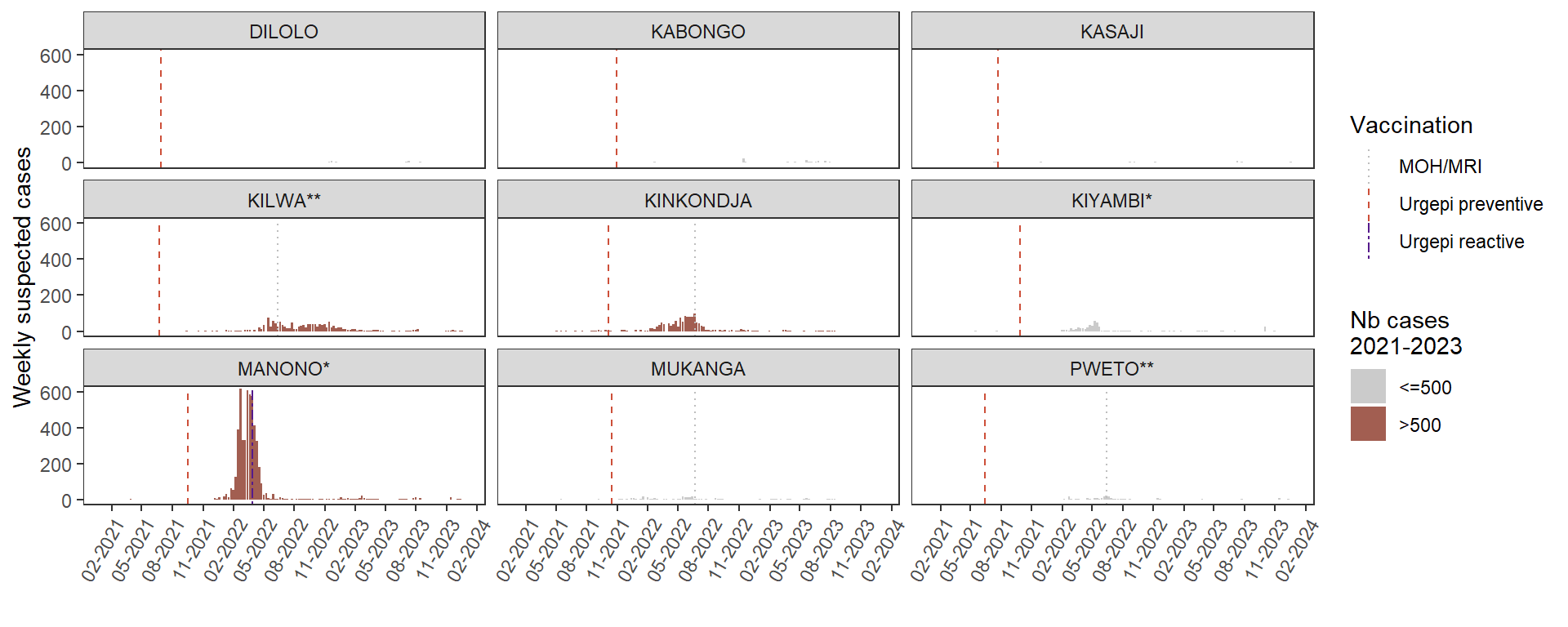

**Figure S7. Number of suspected measles cases notified by health area in Manono, Kinkondja, and Kilwa health zones during 2021-2023.**

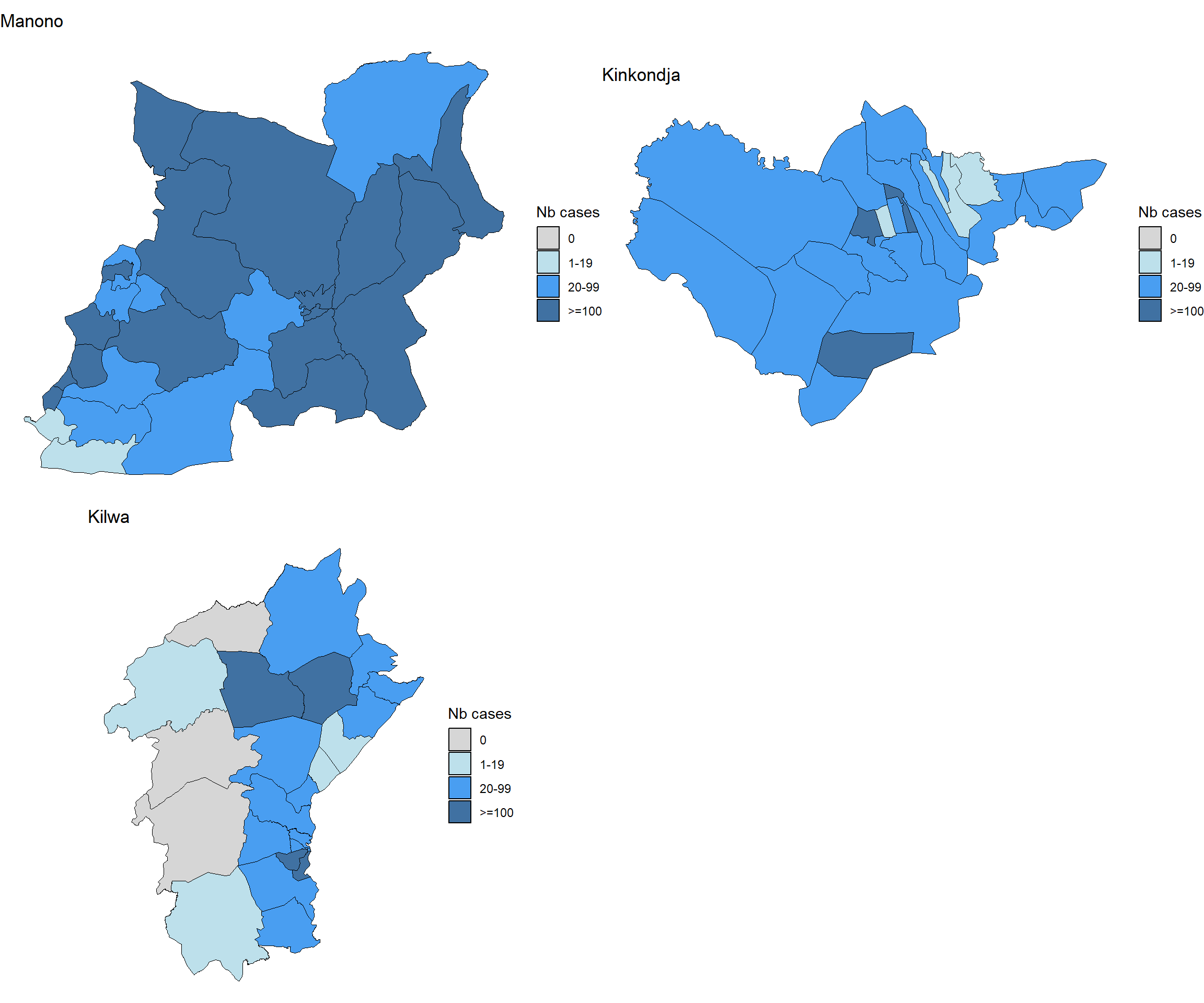

3. **Alert prioritization algorithm and additional figures**

**Table S2: Alert prioritization algorithm.**

| **Criteria** | **ZS to prioritize** | **Score** |
| --- | --- | --- |
| **Epidemiological tendency** | | |
| Biological confirmation | With biological confirmation (required for intervention) | majority of samples negative = 0; biological confirmation in progress / not yet performed = 1; biological confirmation available = 2 |
| Number of suspected cases in the last 3 weeks | With higher number of cases | <10 cases = 0; 10-34 cases =1; ≥35 cases =2 |
| Case number tendency | With an upward trend in the number of cases | Decreasing = 0; stable = 1 ; increasing =2 |
| Case fatality (CFR) last 3 weeks | With a higher lethality | Low or not estimated* (<1%) = 0; high (≥1% - <3%) = 1; very high (≥3%) = 2 |
| **Epidemiological context** | | |
| Priority health zone | Identified as a priority ZS | Non-priority = 0; Priority = 2 |
| Date and size of the last epidemic | No major recent epidemic (within the last 2 years) | Large epidemic  (Attack rate (AR) ≥30/10 000 for 2 years) = 0;  Small epidemic (AR ≥10 & <30 /10 000 for 2 years) = 1;  No epidemic (AR <10/10 000 for 2 years) = 2 |
| Immunization coverage (independent estimate - by survey) | With low vaccination coverage | CV ≥95% = 0; ≥80-<95% = 1; <80 or unknown = 2 |
| Neighbouring health zone in epidemic | With one or more neighbouring health zone in epidemic | No neighbouring health zone in epidemic = 0; 1 ZS in epidemic = 1; >1 ZS in epidemic = 2 |

The maximum alert scores up to 4 weeks after the first week with 20 notified cases (or at any time if 20 cases were never reached) ranged from 6-15 with a median of 12 and IQR of 9-13. Nine health zones reached a higher score up to 8 weeks compared to up to 4 weeks after the first week of 20 notified cases. The probability of large measles epidemics (p=0.02) increased significantly with increasing alert score, although the trend was less pronounced than for alert scores within the 8 weeks window (Figure S8). The increase in median AR was not statistically significant (p=0.17).

**Figure S8 Attack rate per 100 000 (A) and probability of large epidemics (≥500 cases) (B) by alert score up to 4 weeks after the weekly threshold of 20 cases was first reached.** For health zones without notification of ≥20 cases per week the maximum score was considered.

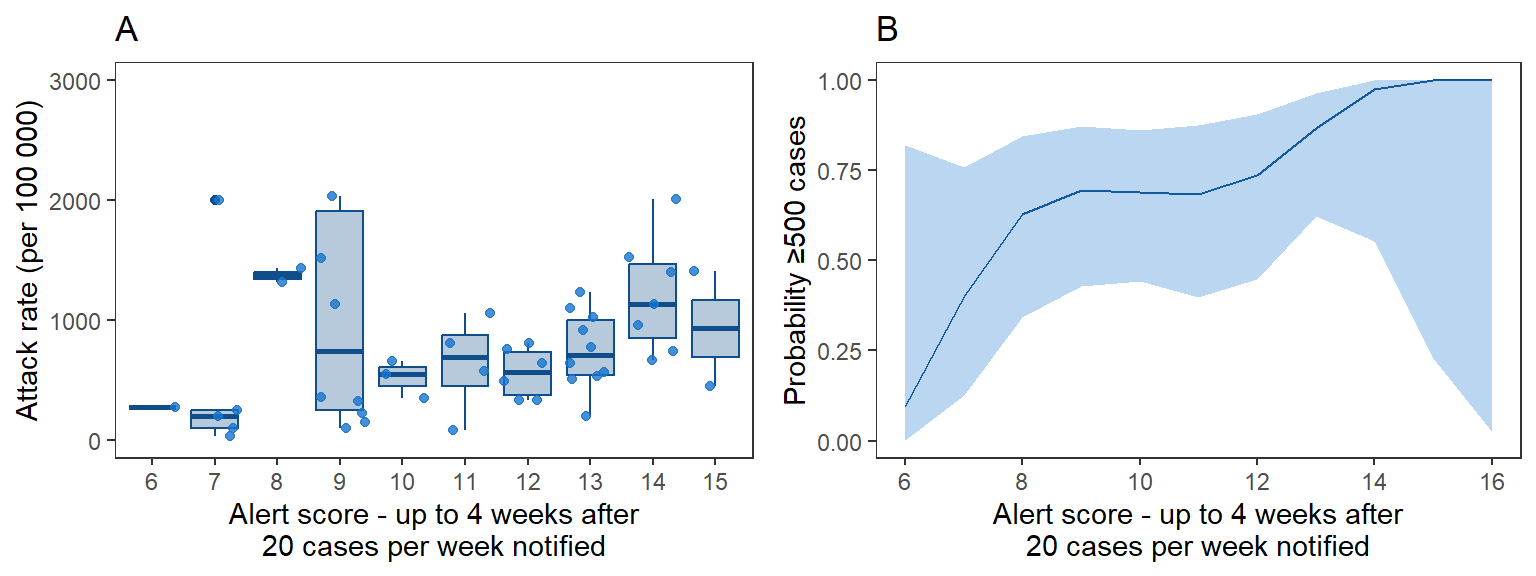

4. **Impact evaluation of reactive vaccination – additional figures**

**Figure S9 Number of cases (A) and attack rate per 100 000 (B) by delay of reactive vaccination since first notification of 20 cases per week.**

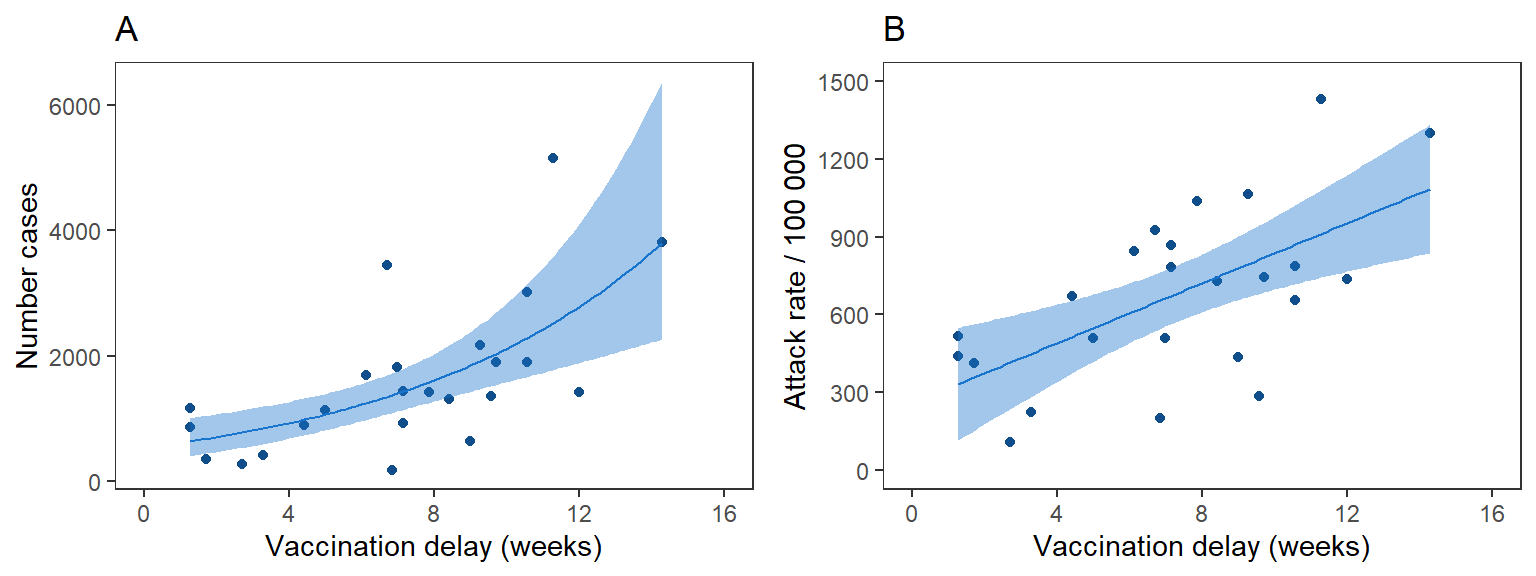
